# Examining the Influence of the Budget Formulation Structures and Processes on the Efficiency of County Health Systems in Kenya

**DOI:** 10.1101/2022.07.17.22277100

**Authors:** Anita Musiega, Benjamin Tsofa, Lizah Nyawira, Rebecca G Njuguna, Joshua Munywoki, Kara Hanson, Andrew Mulwa, Sassy Molyneux, Isabel Maina, Charles Normand, Julie Jemutai, Edwine Barasa

## Abstract

**Introduction:** Public Finance Management (PFM) processes guide the translation of government resources to services and determine health system efficiency. PFM processes are implemented within the budget cycle which entails the formulation, execution, and evaluation of government budgets. We examined how the budget formulation structure and processes influence health system efficiency at the county level in Kenya.

**Methods:** We conducted a mixed methods case study using counties classified as relatively efficient (n=2) and relatively inefficient (n=2) as our cases. We collected qualitative data through document reviews, and in-depth interviews (n=70). We collected quantitative data from secondary sources, including budgets and budget reports. We analyzed qualitative data using the thematic approach and carried out descriptive analyses on quantitative data.

**Results:** Budget ceilings were historically allocated, insufficient, late, or not availed at all. This led to development of budgets that were unresponsive to health system needs. Counties developed both programme-based and line budgets with line budgets as the functional budgets. Line budgets limited accountability and flexibility to reallocate resources. County health funds were fragmented resulting in duplications and wastage. Limited stakeholder participation compromised priority setting and social accountability. Priority setting that was not evidence-informed limited efficiency. Finally, budget changes at the budget approval process compromised alignment of plans to budgets.

**Conclusion:** This study has highlighted six aspects of the budget formulation process in Kenyan counties that ought to be strengthened to enhance health system efficiency: budget ceilings, budget structure, participatory budget formulation, pooling of health funds, priority setting processes and the budget approval process.

**Highlights:**

- Late and Insufficient budget ceilings lead to development of poorly formulated budgets
- Poorly developed and unused programme-based budgets limit health system performance
- Fragmented health system funding results in duplication and wastage
- Limited stakeholder involvement compromised priority setting and accountability

## Introduction

The effectiveness and alignment of public finance management (PFM) processes to health system goals are critical determinants of health system performance and the attainment of universal health coverage (UHC) ^1^. PFM refers to the legal and policy framework, systems, and processes that countries use to mobilize, allocate, spend, and account for public funds ^2^. PFM processes are implemented within the budget cycle that includes the formulation, approval, execution, and evaluation of government budgets ^2^. PFM processes are aimed at enhancing technical efficiency, allocative efficiency, and fiscal discipline in the use of public financial resources ^3^.

Within the health system, PFM has been identified as a determinant of efficiency ^4^. The efficiency of health systems refers to the extent to which health system objectives are met with available resources. Two forms of efficiency are recognized: allocative and technical efficiency. Allocative efficiency refers to an input-output combination that maximizes outcomes at a given cost ^5^, while technical efficiency on the other hand refers to input minimization for a given output or output maximization for a given input ^5^. Increased efficiency may provide a pathway for the attainment of UHC by increasing fiscal space for health ^6^.

Kenya has implemented various interventions to reform its PFM processes across sectors including health. For example, in line with international best practice, in 2005, Kenya introduced the Medium-Term Expenditure Frameworks (MTEF) to link priorities to activities. The Government of Kenya’s Public Finance Management Act (PFMA) 2012 also came up with multiple reforms for the planning and budgeting process including the introduction of programme-based budgeting (PBB) with the aim of strengthening public sector planning and budgeting. PBB was rolled out at the national level in Financial Year (FY) 2013/14 and at the county level in the 2014/2015 FY ^7^. In 2018, PFM processes at the county were digitized with the implementation of the Integrated Financial Management Information System (IFMIS).

Despite these interventions, some PFM challenges have persisted (7). Various studies have documented PFM experiences and challenges across the health system in Kenya. At the national level, one study documented misalignment in the planning and budgeting processes ^7^. At the facility level, there was limited autonomy of health workers in the budgeting process ^8^ and budgets were neither transparent nor credible ^9^. At the county level there was misalignment of the planning and budgeting processes ^10^ and there were challenges with the tools and guidelines to guide the implementation of the PBB ^11^.

Parallel to the PFM reforms, Kenya devolved its governance arrangements in 2013, creating 47 county governments ^12^. The devolution is characterized by fiscal decentralization, with the county governments receiving funds from national government grants (an equitable share block grant and conditional grants), mobilizing additional funds from local revenue collection, and having responsibility for expenditure and reporting ^12^. Within the health sector, the national government retained the policy and regulation role, while the county governments took up health service delivery, including priority setting and resource allocation for health ^12^.

This paper reports findings of a case study of the influence of PFM processes on health system efficiency at the county level in Kenya. The study is part of a larger study that assessed the level and determinants of health system efficiency at the county level in Kenya. In the first phase of the study, a systematic literature review ^13^ and stakeholder engagement ^14^ were conducted that reported PFM processes as key determinants of health system efficiency globally and at the county level in Kenya, respectively. In the second phase, a quantitative assessment of county level health system efficiency and its determinants identified PFM as one of the key determinants of county level efficiency in Kenya ^15^. In this third phase of the study, we explore in-depth how the factors identified in previous study phases interact with county health system efficiency.

In this paper, we focus on the first step of the budget cycle, the budget formulation stage in the Kenyan health sector. This step results in a document that defines government priorities and sets out the path to meet system objectives ^16^. Within Kenyan counties, the process begins with the release of the budget circular and ends with the passing of the appropriation bill. Weaknesses within the budget formulation undermine subsequent steps of the budget cycle and ultimately affect the budget outcomes ^16^. Understanding how the budget formulation process may affect the efficiency of health systems is, therefore, an important research question. While several studies have described challenges with the budget formulation process in Kenya, these studies have not analyzed how these challenges influence the efficiency of the health system. Understanding how processes within budget formulation affect health system efficiency is important informing interventions to enhance health system efficiency.

## Methods

### Conceptual Framework

We developed a study conceptual framework based on a literature review of the influence of budget formulation processes on health system efficiency. The literature review identified 6 budget formulation factors that influence health system efficiency. These are: 1) budget ceilings, 2) budget structure, 3) participatory budget formulation, 4) pooling of resources, 5) priority setting, and 6) the budget approval process. These six issues were reported to influence the health system directly or by influencing subsequent processes in the budget cycle. Figure 1 below outlines the study’s conceptual framework that guided the development of study tools and analysis of study data.

**Figure 1:**
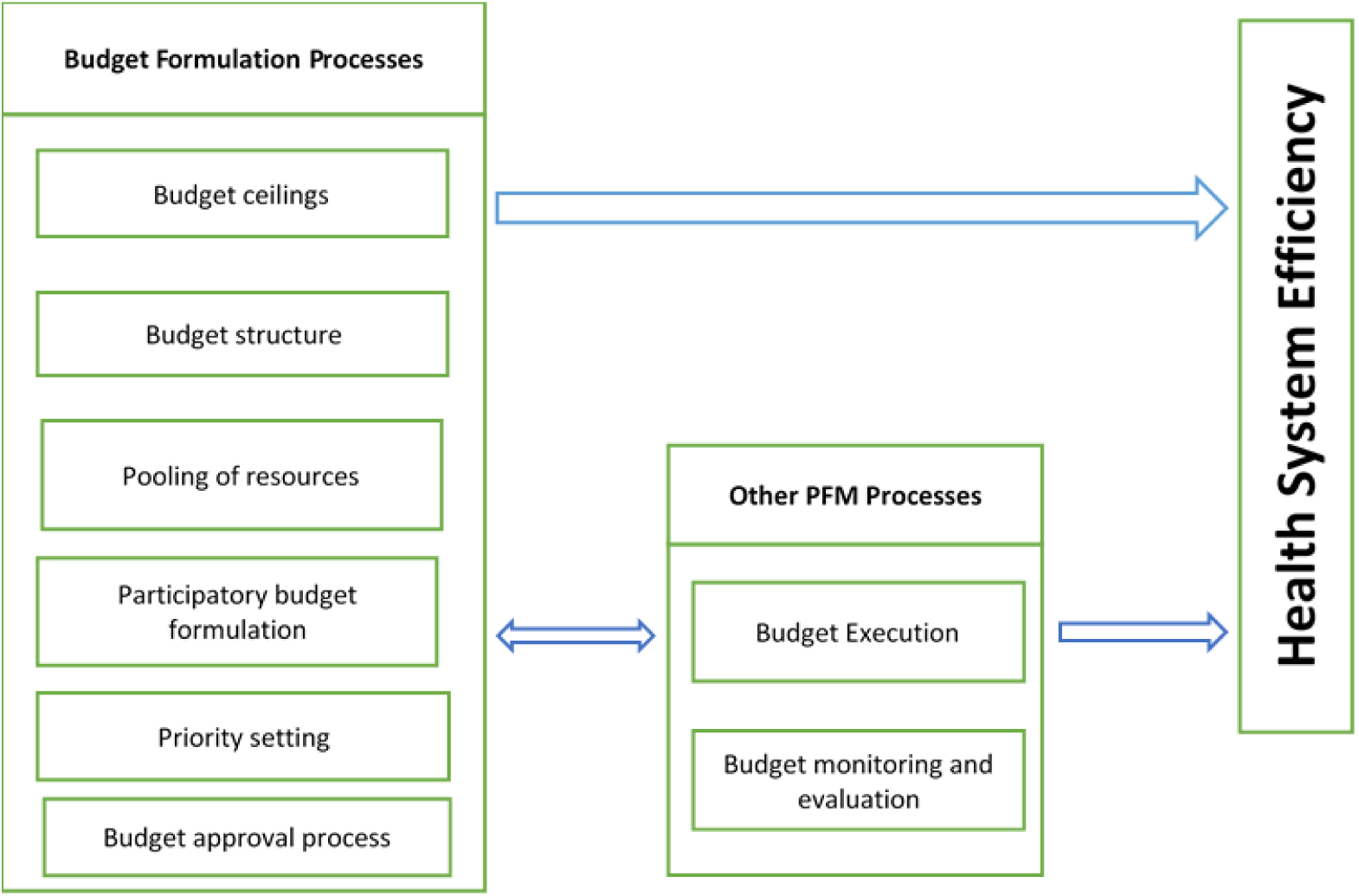
Conceptual Framework.

### Study design

We conducted a concurrent mixed-methods case study. We used quantitative methods to analyse secondary data on budgets and qualitative methods to examine stakeholder perceptions about how budget formulation influences county health system efficiency. The cases for this study were counties – sub national semi autonomous governments in Kenya.

### Study cases

We purposively selected four counties using the level of health system technical efficiency reported in phase two of the broader study ^17^ as a key selection criterion (Table 1). We then took into consideration other county characteristics that were identified by the quantitative efficiency analysis as determinants of efficiency. These are population size and the prevalence of HIV/AIDs. That is, we selected counties with varying population size and HIV prevalence. We included four cases in our case study. To maintain the confidentiality of the respondents involved in the study, we identified the case counties as A, B, C, and D.

**Table 1:**
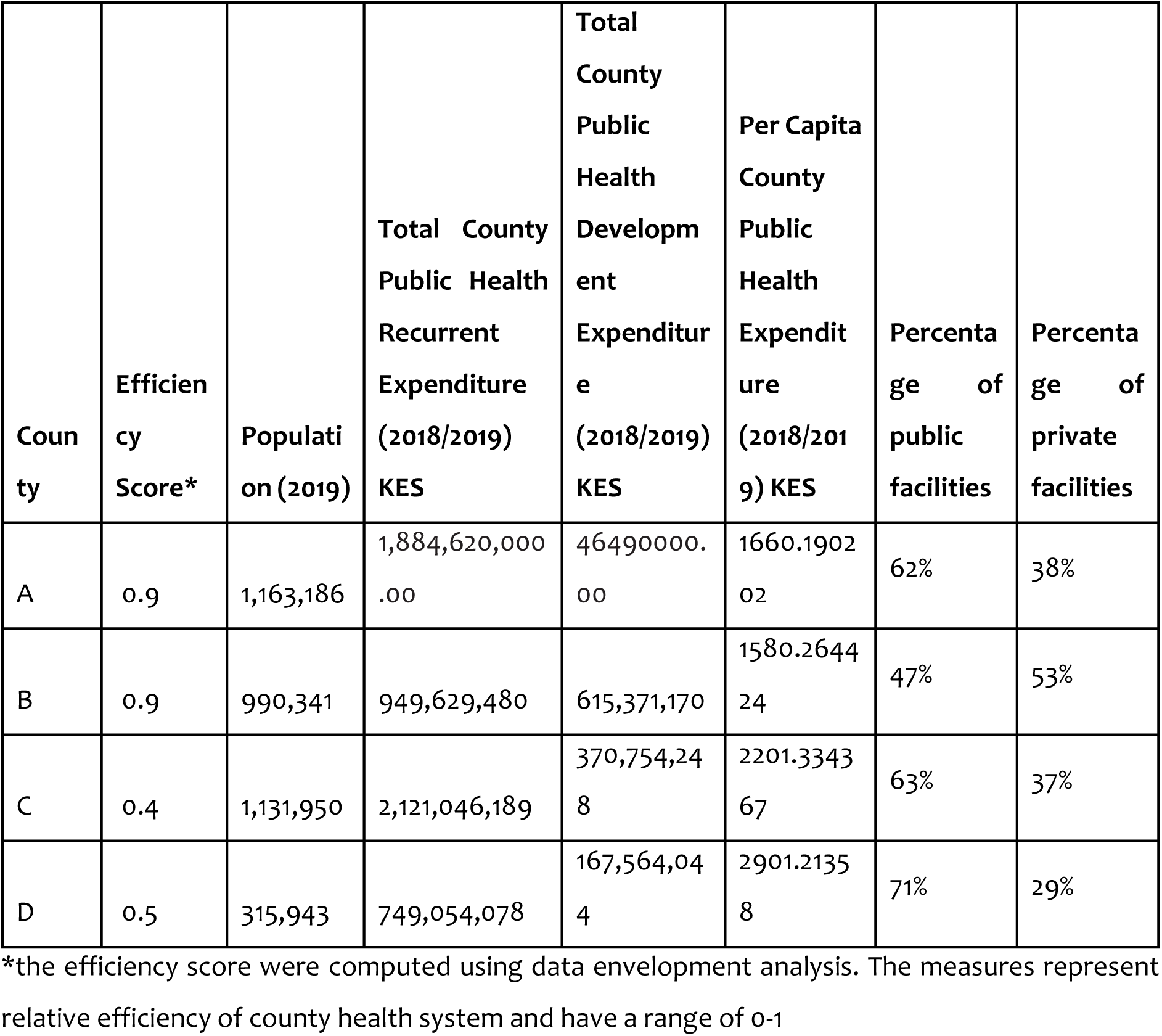
County Characteristics.

### Data Collection

We collected data using a combination of in-depth interviews and document reviews. We collected data between May and October 2021. We purposively selected respondents with knowledge and experience of PFM and health system efficiency. The respondents included facility managers, county department of health officials, county department of finance officials, national treasury officials, ministry of health officials, and donors (Table 2). Two researchers (AM and RK) conducted 70 IDIs in English using semi-structured interview guides developed based on the budget formulation determinants of health system efficiency. All county level IDIs were conducted at the physically while 7 national level IDIs were conducted virtually as per respondents’ preference. The IDIs were audio-recorded using encrypted audio recorders and lasted between 40-90 minutes.

**Table 2:**
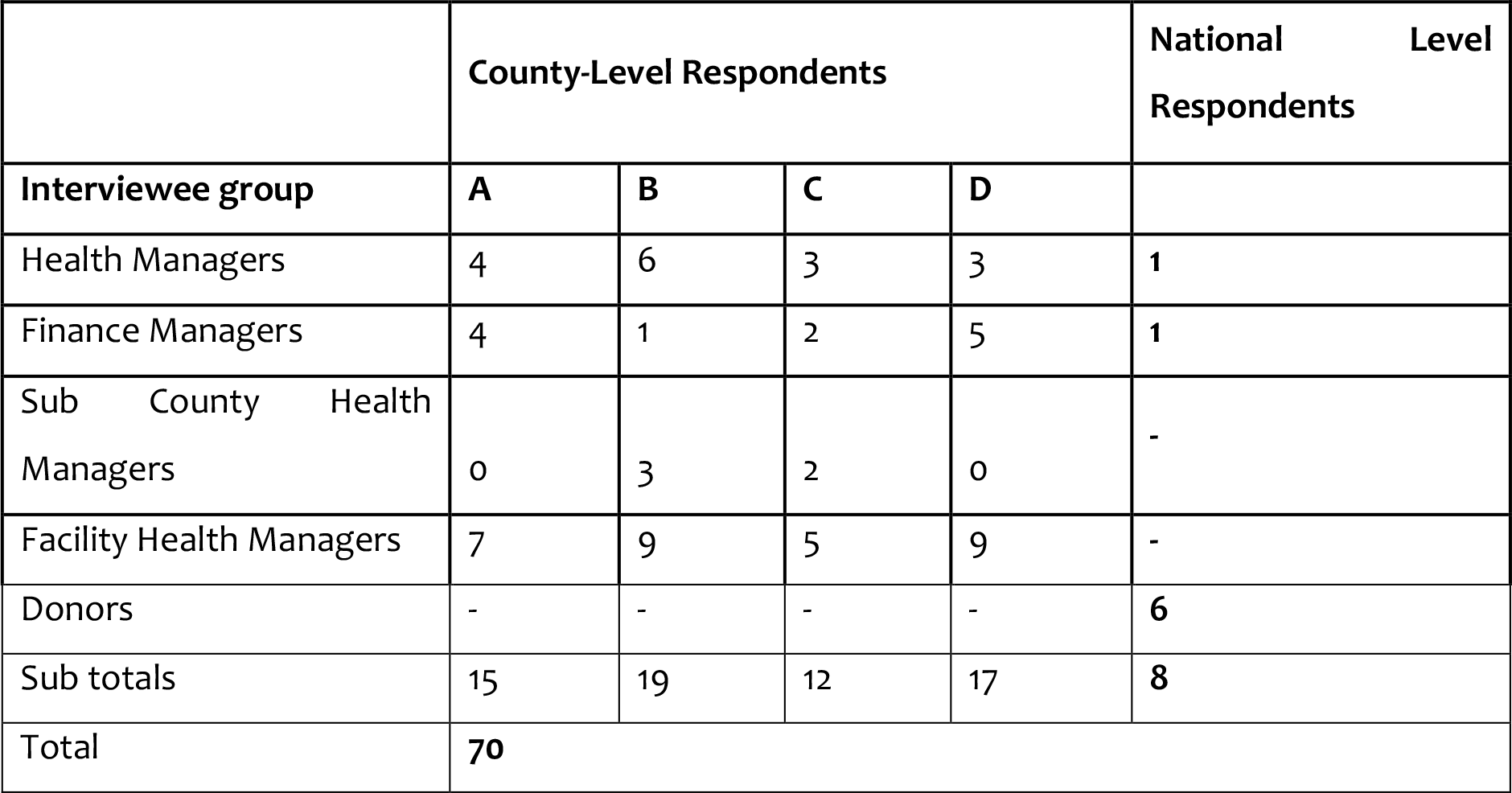
Study Respondents.

We then reviewed budgeting and planning documents for all the four counties for the FY 2018/2019 and national level policy documents on PFM (Table 3).

**Table 3:**
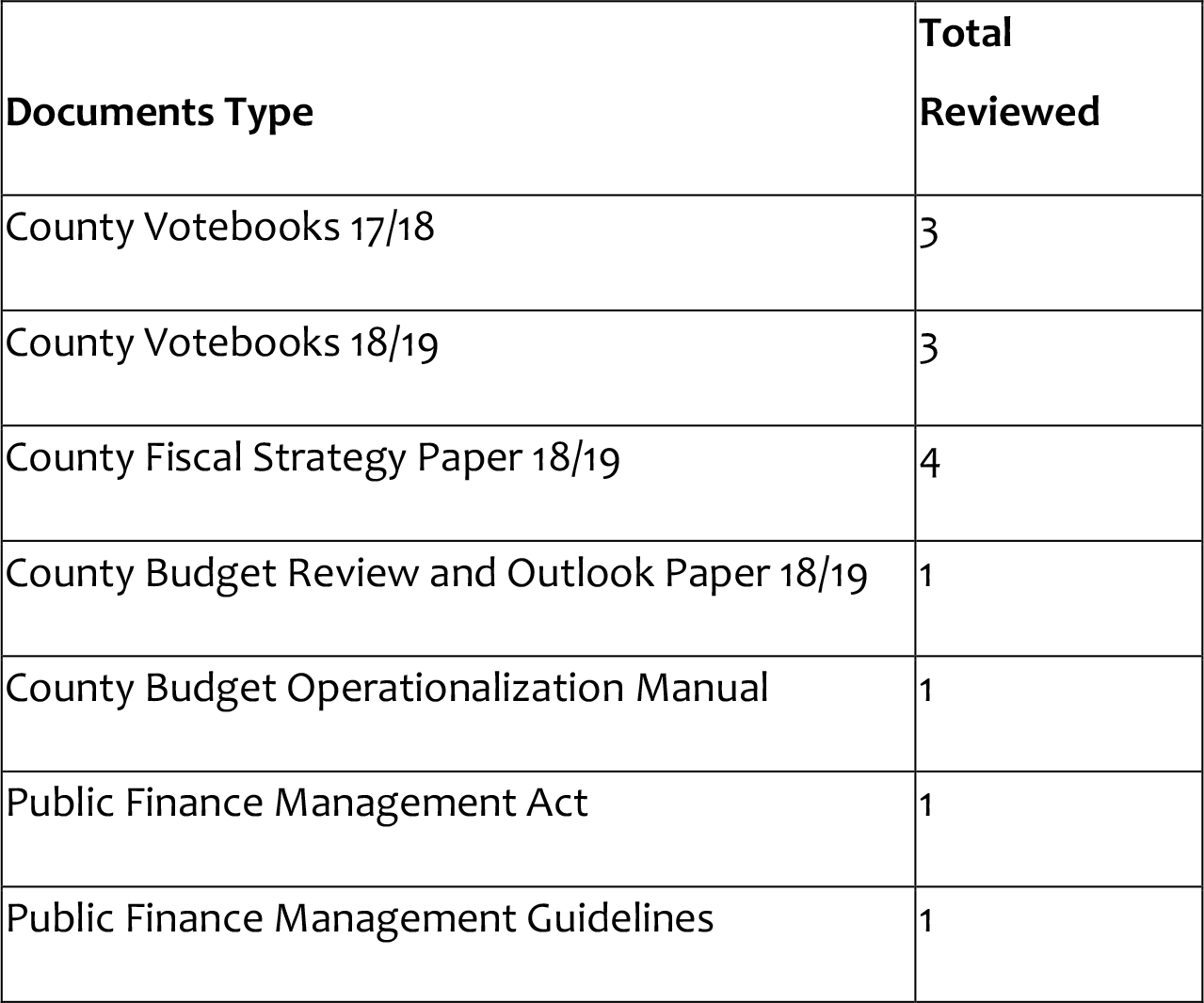
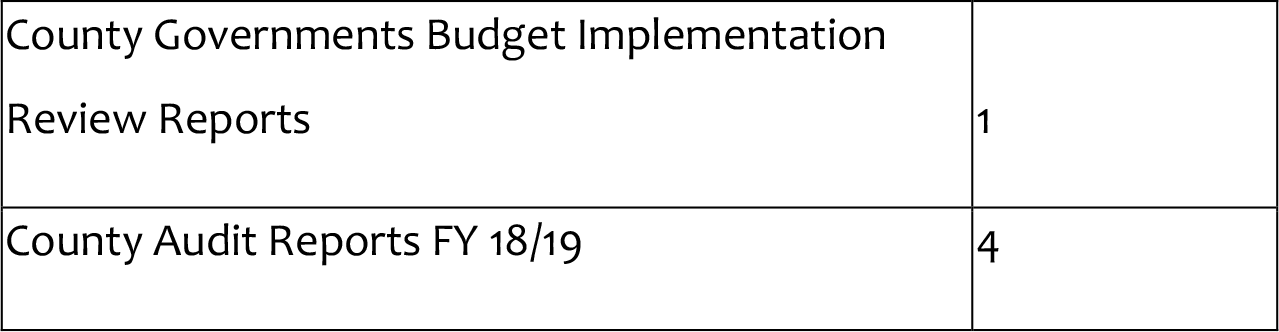
Documents Reviewed.

### Data analysis

Analysis of the data followed a thematic approach. Thematic analysis is a method that guides the identification, organization, description, analysis, and reporting of themes found in a data set ^18^. We immersed ourselves in the data by repeatedly reading through the transcripts. We then developed a thematic framework based on the conceptual framework developed in the literature review, while accommodating emerging themes. Data from the budget process helped to contextualize findings from the review of documents and qualitative interviews. We also held reflexive sessions where we presented initial findings to researchers and policymakers.

## Results

In this section, we first present an overview of the budget formulation process in county health systems in Kenya, then we present the following six aspects of the budget formulation process that we found to influence efficiency: budget ceilings, budget structure, pooling of resources, participatory budget formulation, priority setting and the budget approval process. A summary of our findings per county is outlined in table 4.

**Table 4:**
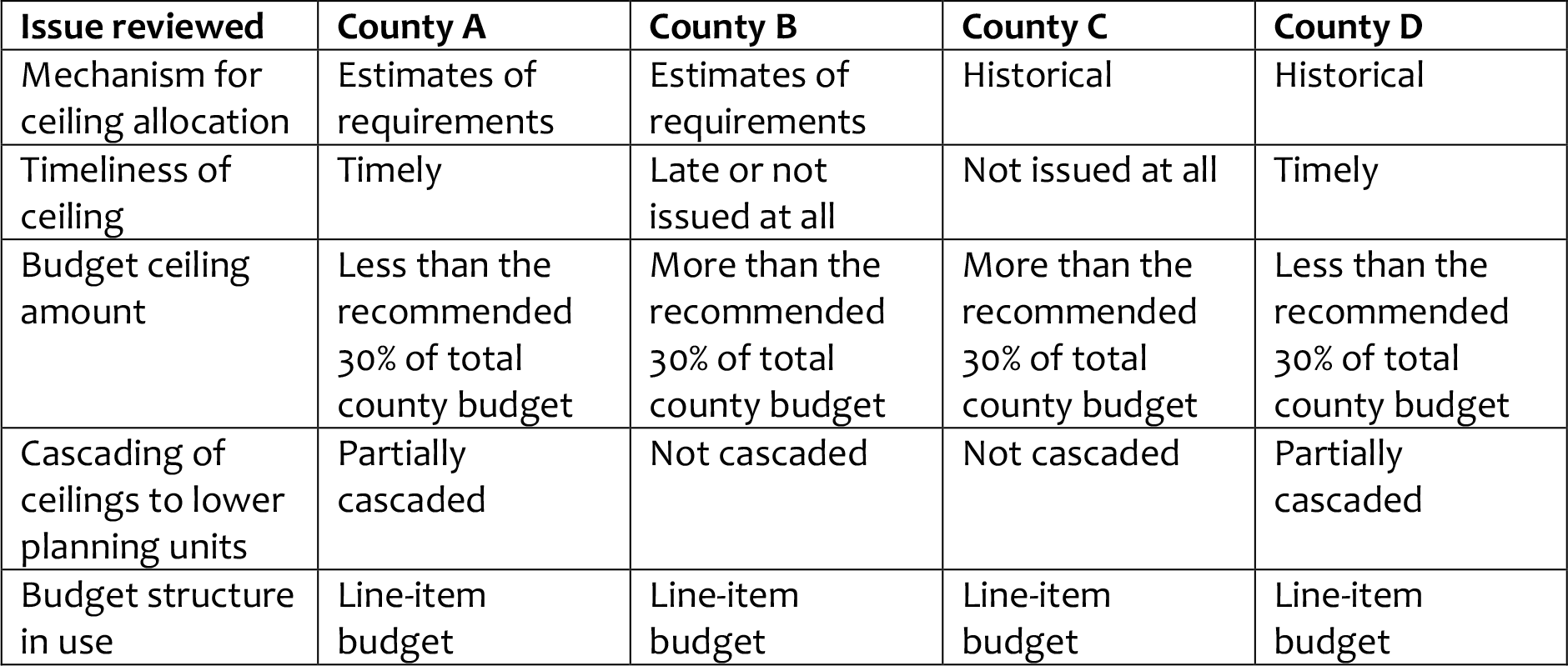
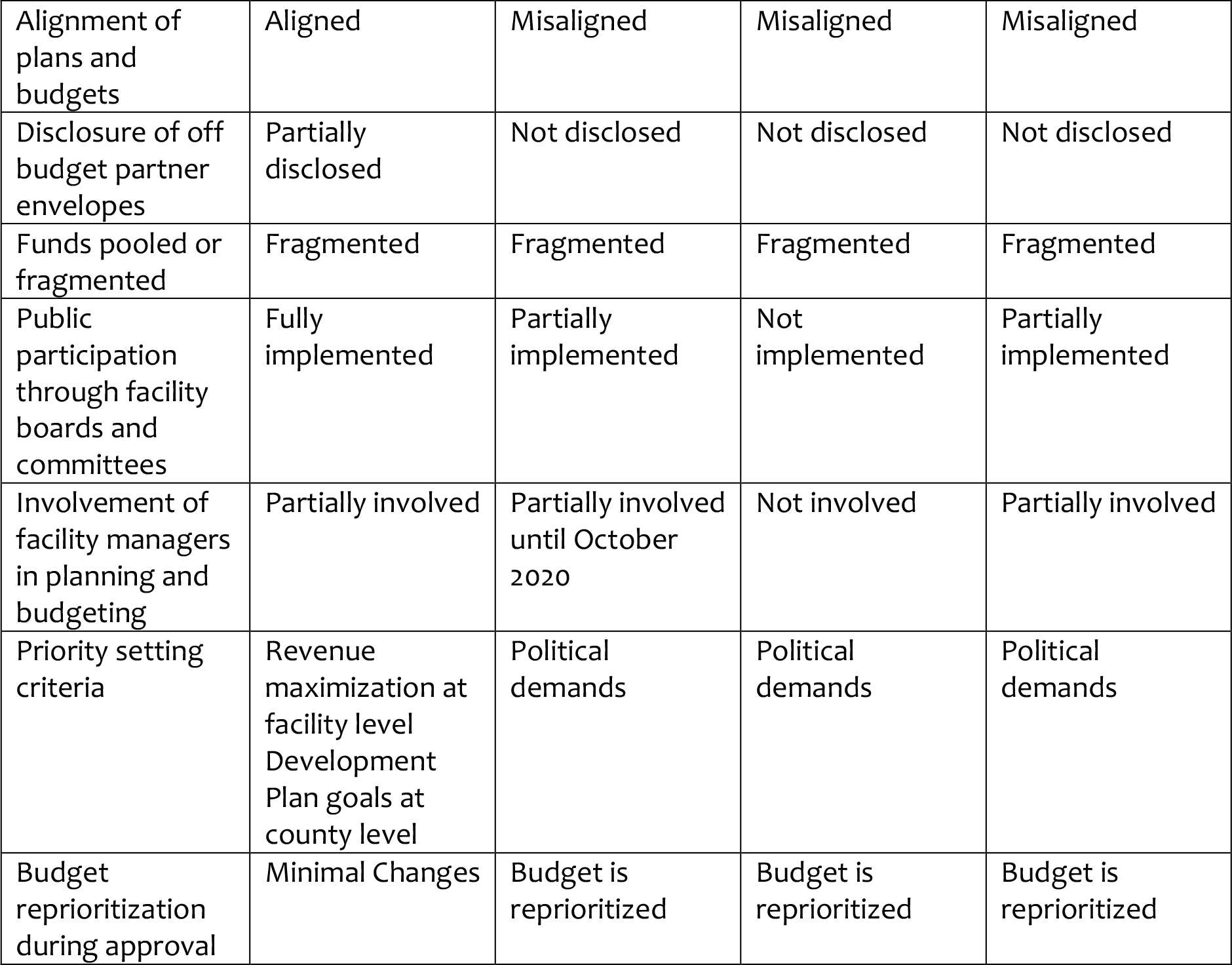
Summary Findings per case study county.

### Overview of the budget formulation process

Kenya’s fiscal year starts on the 1^st^ of July and ends on the 30^th^ of June of the next calendar year. From the policy and legal documents reviewed, the county treasuries release the budget circular by 31^st^ August of every year marking the beginning of the budget formulation process. The budget circular contains key activities and deadlines for the budget process and guidelines for preparing the MTEF budget. The department of finance is also required to submit the Annual Development Plan (ADP) to the County Assembly (CA) and a copy to the commission on revenue allocation by 1^st^ September. The county ADP consolidates sector/departmental ADPs. The department of finance then prepares the County Budget Review and Outlook Paper (CBROP) which is then submitted to the county executive committee (CEC) and CA by 30^th^ September. The CBROP should be published by November. The CBROP assesses the performance of the previous financial year, and makes projections, including proposed budget ceilings, for the next financial year. The CBROP is ideally to incorporate the findings of the departmental Annual Performance Review (APR).

Thereafter, the various sectors, through the Sector Working Groups (SWG) in the county are to prepare the MTEF budgets which identifies priorities in the medium term (3years). The departments should then hold sector hearings where they incorporate public views in the MTEF, thereafter submit it to the department of finance as the final MTEF. By 28^th^ February, the department of finance develops the County Fiscal Strategy Paper (CFSP) to the CA. The CFSP contains the final indicative budget ceilings to the department. The CA is to approve the CFSP by 14^th^ March. Thereafter, this is released to departments who are to develop budgets based on the ceilings and provide proposed budget estimates to the department of finance. The department of finance should then compile the proposed estimates and submit them to the CA together with the supporting documents by 30^th^ April.

Between May and June, the CA budget appropriation committee should then conduct public hearings of the proposed estimates. Thereafter the CA is required to approve the estimates by 30^th^ June, becoming the approved budget (Figure 2)

**Figure 2:**
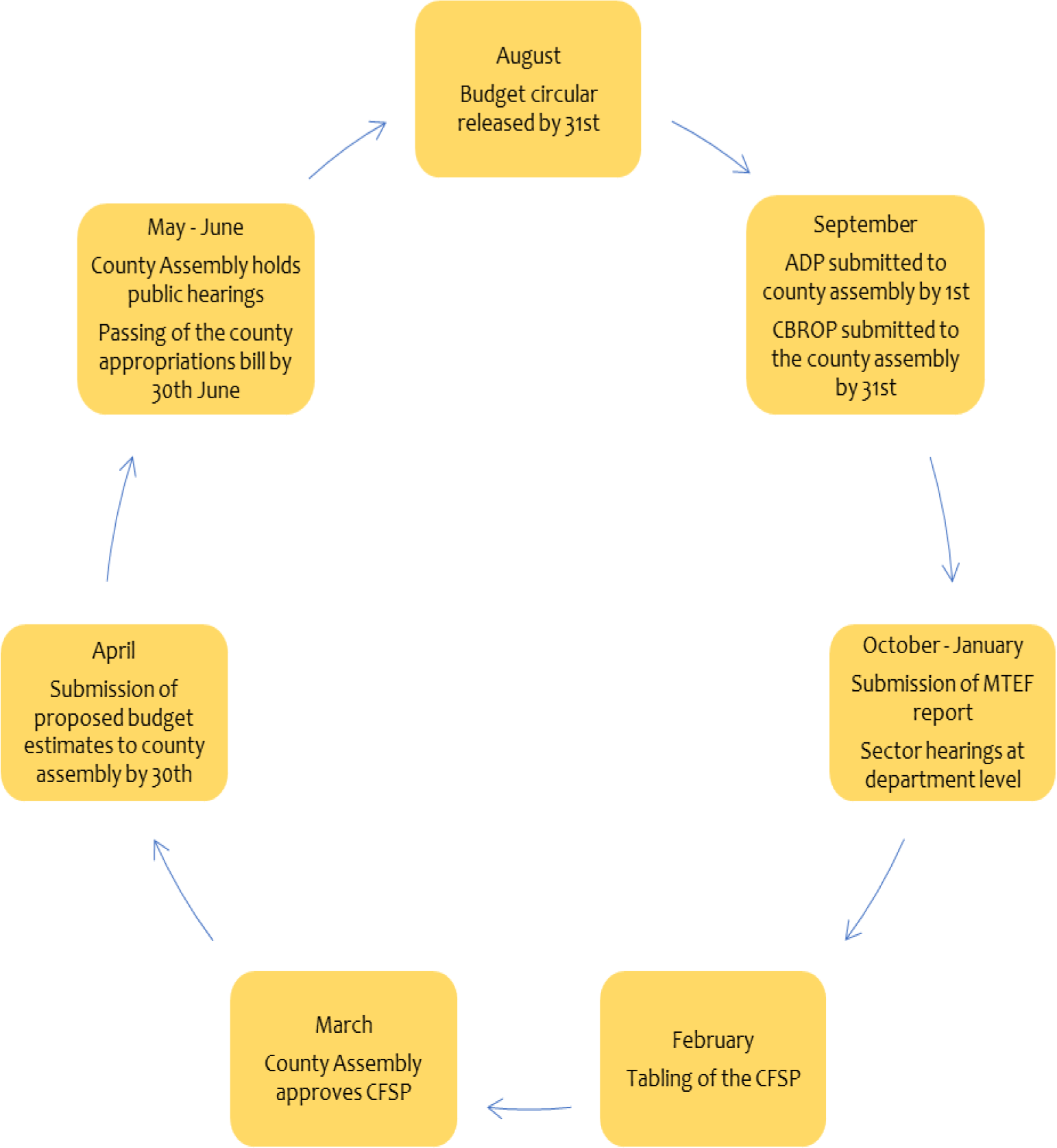
Budget Formulation Process within Counties in Kenya

### Budget ceilings

#### Across the case study counties, there was a variation in how budget ceilings were determined

In counties A and B, ceilings were determined using estimates of financial requirements for health system inputs (e.g. salaries, drugs). However, in counties C and D, ceilings were determined using historical allocations. These were adjusted by a percentage in county C, and by the department budget absorption in county D.

> *“The ceiling is decided based on costs for critical items. These include staff salaries and essential health commodities such as non-pharmaceutics. It also includes projects that have political priority or are ongoing. For instance, in our county, the construction of the teaching and referral hospital is a political priority for the Governor and has been ongoing. This is prioritized in budgeting to avoid stalling of the project”County Finance Manager, County B*

Interview respondents reported that the use of historical allocation to determine budget ceilings limited the budgeting processes’ responsiveness to the changing needs of the county.

> *“Historical budgeting carries forward budget challenges from previous years to subsequent years” County Facility Manager, County C*
>
> *“Any new activities that were not in the previous budget are not included in the new budget and hence are not funded” Facility Health Manager, County D*

#### Some respondents noted that ceilings to CDoHs were availed late or not at all

While ceilings were availed on time in county A and D, they were not availed in county C. Ceilings for county B were either availed late in some financial years or not at all in others. Respondents reported that ceilings were not availed because of bureaucratic inefficiency, or because the county treasury made the final budget on behalf of all the other county departments. Interview respondents across the counties felt that the timeliness of availing of ceilings to the CDoH influenced efficiency in various ways. First, ceilings that were not availed limited effective planning within the health sector and contributed to historical budgeting. For example, in County C, it was reported that in the absence of budget ceilings, the CDoH limited its planning to the previous year’s budget yet the current years resource envelope sometimes varied considerably from the previous years. On the other hand, in County B, it was reported that ceilings that were not availed shifted the decision-making power on priorities to the county treasury and hence reducing the CDoH’s autonomy over health sector resource allocation.

> *“We did not know the ceiling so we would limit ourselves to the previous figure. As a result, we would fail to budget for some things. And, even after restricting ourselves to that previous figure we did not know what ended up in the budget, it is finance that would decide what to include in the final budget because they had the real ceiling” County Health Manager, County B*

Secondly, it was felt that late ceilings did not give time for adequate consultation hence key inputs and information were missed out on the budget. For example, in County B, respondents from the CDoH reported that they only had one week to compile a budget, as a result, only two people were involved in determining the needs of the whole county. Because of the time and the few people involved, they left out important issues in the budget. In county A, the county tried as much as possible to involve all relevant stakeholders.

> *“Budgeting has become a two-people show involving me and Dr. XXXX. We stay up late trying to budget for the county. If we had more time, we would budget as a team. A bigger team would bring diverse views and ensure nothing is left out” County Health Manager County B*

Thirdly, interview respondents reported that late budgeting limited the use of evidence in the budgeting process and contributed to historical budgeting. For example, in County B, it was reported that the budgets for previous years were replicated because they did not have time to make alternative considerations.

> *“One week is the longest we’ve ever had, so if you look at our previous budgets, they are almost the same because you don’t have time to think a lot” County Health Manager County B*

In counties where ceilings were availed on time, respondents reported that this facilitated the timely processing of the budgets and ultimately, the timely release of funds for activities.

> *“The budgeting cycle is interrelated, with clearly set timelines. The moment you are late in one process, the other processes are affected. So timeliness has enabled the smooth flow of the timelines and the final implementation of the budget” County Finance Manager County D*

Finally, interview respondents reported that timely ceilings allowed for the early identification of budgetary gaps. This gave the county health management team time to source alternative funds to bridge the budget gaps.

> *“Spending caps inform us on the budget limits. Thereafter, we can realize budgetary funding gaps. We then work with donors to plug in the deficits” Facility Health Manager County A*

#### Respondents in all the counties at both county and facility levels noted that budget ceilings were not sufficient

The intergovernmental participation agreement recommends that counties allocate a minimum of 30% of the county budget to the CDoH. Document reviews show that allocation levels to the health sector varied across counties, with counties B and C exceeding the recommended minimum, while counties A and D allocated below the recommended minimum (Table 5).

**Table 5:**
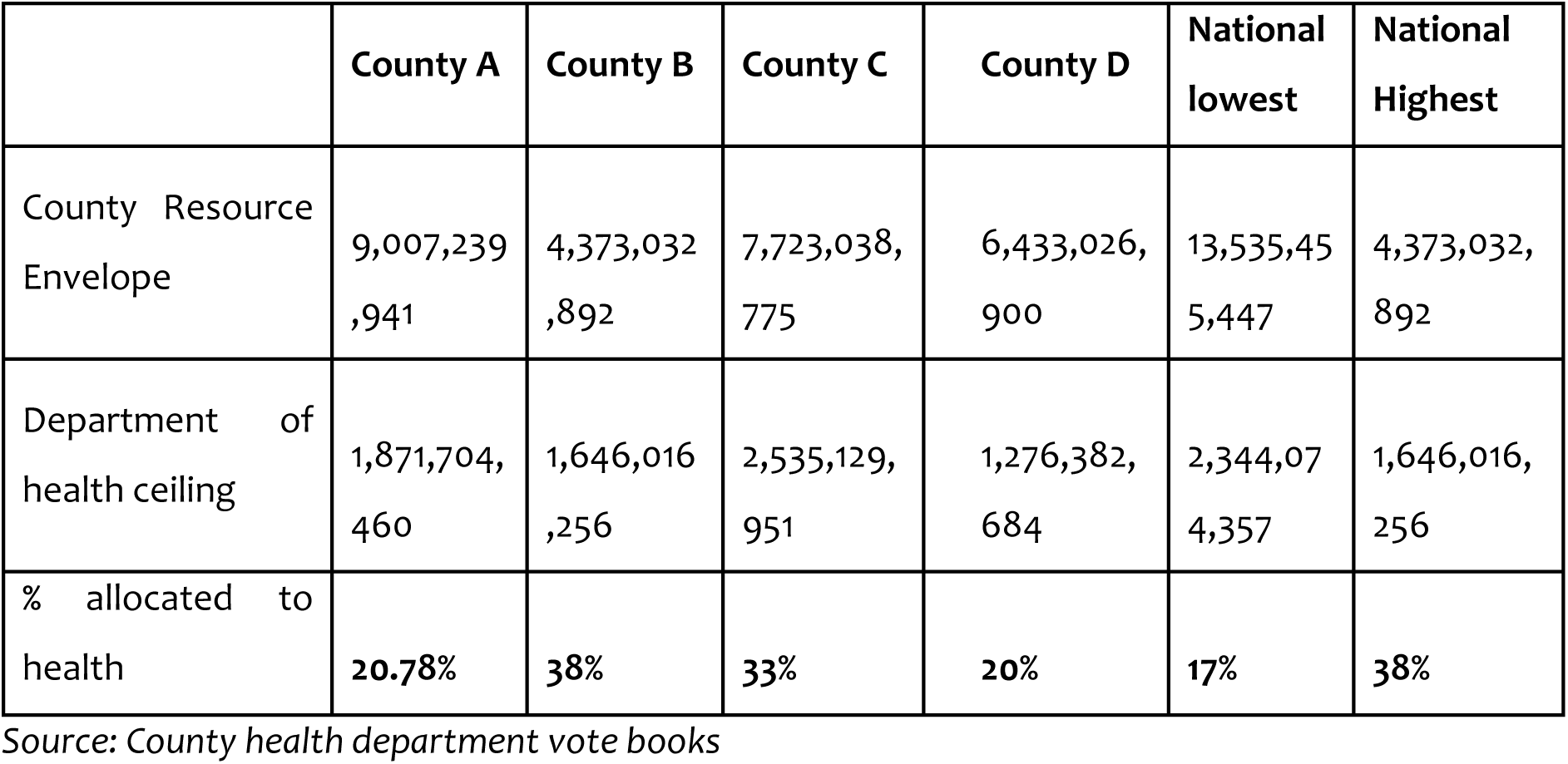
Total County Allocation to Health.

Interview respondents felt that insufficient budget ceilings affected the efficiency of county health systems because some activities had to be left out of the budgets and were not implemented.

> *“Sometimes we exhaust the allocation, and we have to operate on credit. Hence, we must limit the services. For instance, maternal referrals require a fueled ambulance and an allowance for the accompanying nurse. If funds are limited, patients will have to pay for the services out of pocket. But often clients are not prepared” Facility Health Manager, County C*

#### Budget ceilings were not cascaded to the lowest planning unit

despite this being a requirement of the county budget operationalization manual. For example, in counties A and D, only ceilings for donor funds were cascaded down to level 2 and 3 (primary level) facilities while in counties B and C no ceilings were cascaded down to facilities. Some respondents from the County Health Management Team (CHMT) felt that it was difficult to cascade ceilings because the ceilings were not sufficient.

The CDoH failure to cascade ceilings to the frontline was felt to influence health system efficiency in various ways. First, as there was no ceiling, frontline workers made unrealistic plans which were either partially funded or not funded at all. Also, it was the CDoH rather than the facilities that decided what was funded, which limited frontline autonomy.

> *“We are not given a budget ceiling; we budget based on our needs. As a result, budgets are not honored or less than 50% is honored hence we cannot implement our plans. For example, we do not get health products in the required quantities. When we exhaust the available products, patients have to purchase goods and services out of pocket” Facility Health Manager County C*

Second, it was felt that failure to cascade down ceilings resulted in ad-hoc short-term budgeting at the health facilities. Facilities only made budgets when they received funds. This limited long term planning which was geared towards achieving targets and goals.

> *“As facilities, we are using a line-item budget, because, a program-based budget would only be appropriate if we have an annual budget, which does not happen in health. Because we are not sure when the money will hit our account. So we only budget when the money hits the account” Facility Health Manager County C*

### Budget Structure

The Government of Kenya (Kenyan government) budgeting legal and policy framework requires that entities develop programme-based budgets (PBBs). PBBs link resources to programmes, activities, and indicators thus enhancing efficiency. During the budget formulation process, the CDOH made PBBs. Funds for programmes were then broken down into sub-programmes and line items. Following approval of the budget, and once the budget was uploaded on Integrated Financial Management System (IFMIS), the finance office then generated a line-item budget which was then issued to the department for implementation. Further, respondents in all the four selected counties noted that health facilities developed line-item budgets.

The formulation of line-item budgets was felt to influence health system efficiency in various ways. First, it was felt that the budget was reduced to a document that financed inputs rather than outcomes. The budgets were made to mitigate short term input crises rather than achieve sector goals and objectives

> *“The challenge with line budgets is we ignore some activities. It is a reactive budget, when we run out of food we buy food, same applies to fuel. Programme-based budgeting is more inclusive, nothing is left behind. But even if we make programme budgets, we will never get the funds” Facility Health Manager County C*

Second, respondents felt that line-item budgets limited accountability for service delivery outputs and outcomes. It made it difficult to link the budget to programmes that were implemented. The line budget was a financial accounting document that explained how resources were expensed, however, it did not provide an opportunity to evaluate indicators.

> *“Itemized budgets are majorly an accounting document which the accounting department can use to account for e money, but for us now, we cannot link it to the indicators*.*” County Health Manager, County D*

#### County health budgets were not aligned to plans and targets

Interview respondents felt that misaligned budgets and plans affected efficiency in various ways. First, it was felt that performance indicators were either not included in the PBB or, where they were included, they were linked to intermediary rather than final health system outputs (Figure 3). This limited the benefits of the PBB process thereby limiting efficiency. For example, it limited accountability for outputs and outcomes as resources were allocated but there were no tangible outputs or outcomes attached to it.

**Figure 3:**
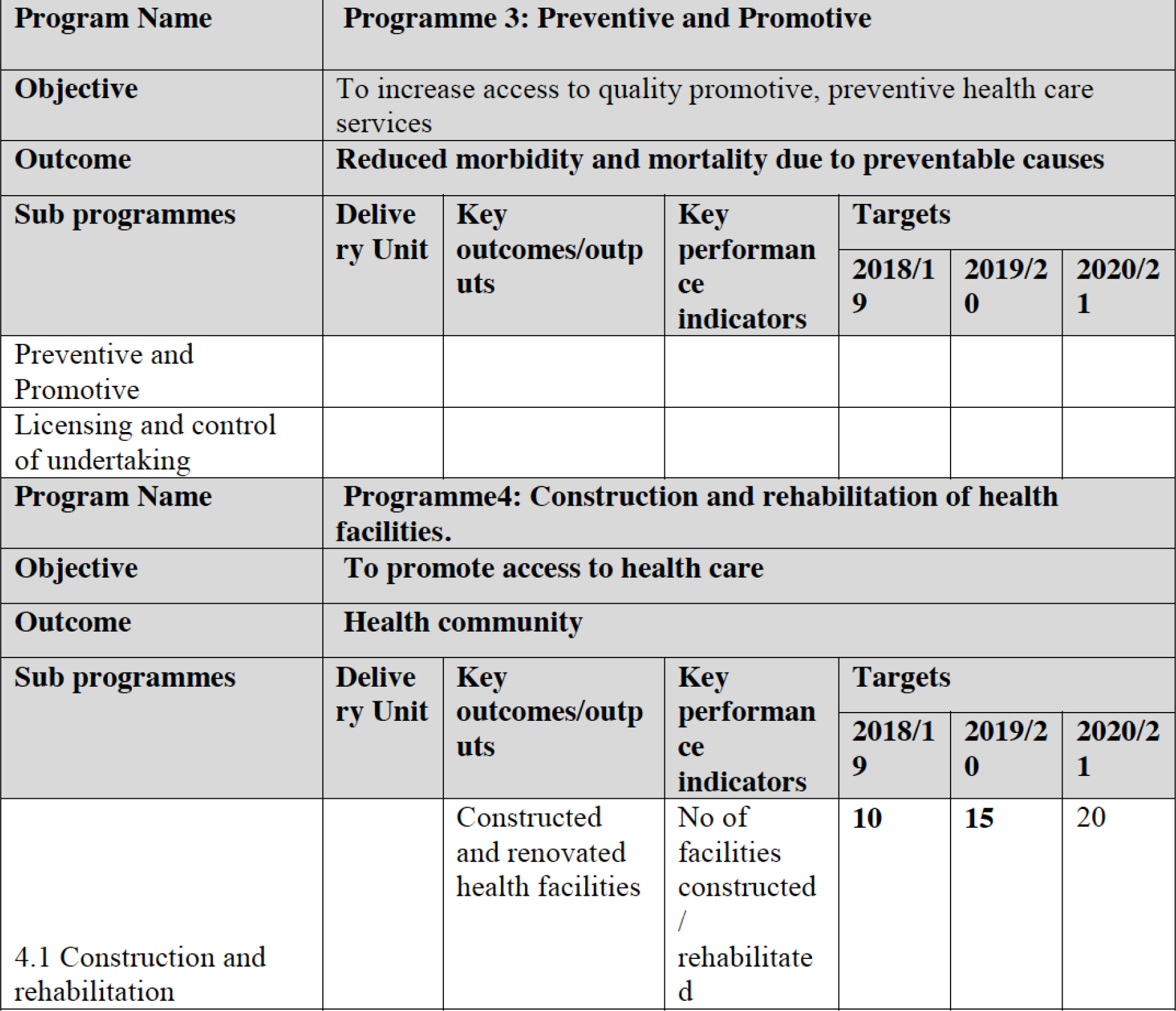
Sample county programme-based budget indicating outputs/outcomes.

Secondly, respondents felt that the misalignment of plans with budgets led to a mismatch in the inputs available to the health system leading to wastage of resources. The input combination was targeted to maximize expenditure contained in the budget rather than outcomes which were in the plan. This resulted in an input mix that could not deliver the intended outcomes. For example, in county C, the government provided cervical cancer vaccines but did not budget for resources to conduct vaccination within schools to ensure vaccines reach the population. As a result, the vaccines expired.

> *“At the moment, we have 48 closed laboratories, because of staff [shortage]. The infrastructure and equipment is lying there unutilized because we don’t have staff yet we present our needs every year in the annual work plan” County Health Manager County C*

Third, respondents felt that misalignment between plans and budgets had a negative impact on health outcomes.

> *“Because XXXX borders the game park, we have many fatal snake bites. So in our annual work plan, we report the number of deaths and plan to purchase anti-snake venom. We have been putting this in our annual work plan for the past three years, but the county has never provided the anti-venom” Sub County Health Manager County C*

Finally, misalignment in plans and budgets was felt to compromise accountability for results. Monitoring and evaluation of budgets against indicators could not be accomplished.

> *“We can’t evaluate indicators because the activities of the indicators do not go hand in hand with the financing. The only indicator that you can evaluate are the donor funded ones. If donors support reproductive maternal and child health, they are very clear on the specific indicators that they chase alongside that budget*.*” County Health Manager County D*

### Pooling of resources

#### Respondents noted that one key challenge for pooling across the various funding streams was the donors’ failure to disclose their budgets

In all four counties, only two donors provided on-budget support to the counties and hence disclosed their resource envelopes (the World Bank and Danish International Development Agency (DANIDA)). In county C, some respondents felt that the failure to disclose resource envelopes was because of fears of interference from political leaders. It was reported that if donors were to disclose their resource envelopes, then local leaders such as the MCAs would want to skew the allocation of resources to their advantage rather than the health system’s advantage. Another reason given by donors was that the county expected information from them, but the counties were not forthcoming with information, they stressed the need for a two-sided rather than one-sided relationship.

Failure to provide budgets was felt to affect health system efficiency in multiple ways. First, it was felt to undermine the priority setting process, as departments were either uncertain about allocating activities to donors who did not disclose their envelopes, or they made incorrect assumptions about the level and support of donors thereby leaving out key activities from their budget.

> *“Undisclosed envelopes are a problem because we make lots of assumptions when budgeting. For example, we do not budget for HIV and TB with the assumption that they are covered by a donor or the national government. But sometimes not everything is done yet on the budget we omitted it. Besides there is a danger of duplication, this is worse because that money could have funded other activities” County Health Manager County B*

Second, it was felt that it led to duplication of activities. This led to the wastage of resources, yet some key activities remained unfunded. For example, donors provided commodities that health facilities had already purchased, and they had no option but to accept the commodities. Also, donors facilitated outreach activities that facilities had already included in their budgets. While the facilities appreciated the input, they had to go through a long process to reallocate the budgeted funds.

Third, interview respondents stated that they were unaware of the donor support and this limited sustainability of the donor implemented activities. For example, in county D, it was reported that while the county received a lot of support, the county government was unaware of this as the donors did not disclose their envelopes.

> *“Donors provide a lot of support, but this is not felt because they do not disclose their contribution. For instance, within the nutrition department, organizations like XXX and XXX support us with nutrition commodities, they spend a lot of money but the county is unaware of the value. If the donors pulled out today, the county may not be able to take over because they lacked full information” County Health Manager County D*

#### Funding to the CDoH was through fragmented channels

The funding sources for the four CDoHS can be categorized into three: exchequer allocations, county own source revenue, and donor support. The exchequer support came in various forms. These included routine allocations (comprised of development budget and recurrent budgets) and conditional grants. The conditional grants were mainly for user fee foregone by primary health centers and grants to county tertiary health (level 5) facilities. None of the four counties included in the study received a county tertiary health facility grant. Own source revenue, on the other hand, was funds generated from various activities for the health sector. These included user fees, and insurance funds such as Linda mama (an MOH insurance for all pregnant mothers in Kenya), Edu Afya (MOH insurance for all Kenyan school children), National Health Insurance Fund (NHIF) capitation and fee for service reimbursements. Donor support came through either on-budget support or off-budget support. On budget support was from two sources i) the World Bank *transforming health systems* (THS) funds to the counties in general for improving Reproductive Maternal Neonatal Adolescent and Child Health (RMNCAH), and ii) the Danish DANIDA conditional grant to improve primary healthcare facilities. Off-budget support also came in multiple ways, some through direct financing via cheques to County Health Management Teams (CHMTs) commercial bank accounts, and in-kind support through the provision of inputs.

Fragmented financing of the health system was felt to be affecting health systems efficiency in various ways. First, it was reported that different sources of funds targeting specific areas of implementation led to the fragmented achievement of health system goals. For example, in County D, it was reported that one facility was unable to meet Immunization goals for the Bacillus Calmette Guerin (BCG) vaccine because they did not have a maternity wing. The funds they had could not be used for the construction of a maternity wing as they targeted only immunization activities. In County B also at the county level, the RMNCAH indicators were doing very well as they were supported by the donors, but other areas lagged.

Second, it was felt that fragmented funding sources led to fragmented decision-making for budget priorities. This resulted in duplication of activities and therefore wastage of resources. It also provided a loophole for informal priority setting which led to inefficiencies.

> *“We advise MCAs to have one model facility to serve people across political areas, however, they will hear none of it. Every MCA wants a facility for their area, even if there is a facility within a 1-kilometer radius of the population. The MCAs will go ahead and allocate finances to construct a facility in that location. When the CDoH disputes the allocation, the MCAs reallocate the funds to other departments therefore the health department loses out. Having fragmented development funds for administrative wards is a wrong model, all these resources should be under the CDoH” County Health Manager County B*

Finally, respondents felt that multiple accountability channels for the different resources led to challenges with accountability. For example, in county B, MCAs put support in the very same activities that were supported by donors. Thereafter they did not implement the activities budgeted by the counties, yet they still consumed the resources.

> *“I thought our county is bad, but I am informed that other counties are worse. Some counties own the donor-funded projects, they claim they paid for the project. Of course, we lose out. Someone will gain, someone will be paid. There was no contractor but someone said they did work which was done by someone else so, in essence, it is a double payment but because the processes are sort of independent whatever returns will be made to the donor and whatever returns will be made to the county they may never meet” County Health Manager County B*

While there were several inefficiencies from fragmented funds, interview respondents felt that fragmented financing also helped the health facilities especially when the main source of funds were unreliable. Alternative financing sources such as off-budget donors, Linda Mama and NHIF came in handy.

> *“Other counties are performing. So, it makes us question the priorities of our county. Were it not for MSF providing services at the county hospital* …*it would also be down and under. The county is over-reliant on MSF aid, for things that the county itself should be providing* …*” Facility Health Manager County C*

### Participatory budget formulation

The Kenyan government legislative framework requires a participatory budget process. In the period prior to devolution of healthcare services, the Ministry of Health (MOH) constituted mechanisms for structured community involvement in decision-making within health facilities through health facility management boards and committees. These boards and committees involved community members and frontline health workers in the budget and planning process.

> *“…there shall be openness and accountability, including public participation in financial matters” – Constitution of Kenya 2010*

From the document reviews, however, we found that the devolved government system introduced public participation into the budgeting process through sector working groups (SWG) and public barazas, in addition to the facility committees and boards. Interviews with county-level respondents reported that the health SWGs in most of the counties were not functional.

#### The respondents felt that while the public was involved in budget formulation, their involvement was unstructured

Public participation was felt to influence health system efficiency in various ways. First, it ensured that the health budget was responsive to the needs of the population. For example, in county C, it was reported that the public was keen that all the implemented activities were responsive to their needs.

> *“I have been in this system for 30 years but for the last 10 years, people have been very alert. You cannot go to the community and implement projects that are not in tandem with community expectations, they will reject the project. The days when doctor’s word was final are long gone” County Health Manager, County C*

Second, it was felt that the public was able to lobby for more resources on behalf of the county health department.

> *“they’re [the public] the ones who lobby the county government to bring new projects to our facility” Facility Health Manager County B*

Third, respondents felt that unstructured public participation led to conflicts between provider needs and community needs. For example, in County D, the finance department noted that there were differences between the public and facility needs, and finance had the responsibility to balance the different requests.

> *“There are these public participation forums that the county holds, we as the health workers, rarely attend. They are held when we are at work. Therefore, our views are not factored in, yet as a service provider we know what is needed to improve outcomes”* Facility Manager County D

Respondents noted that before devolution, the MOH had clear structures that defined how the public was involved in the budgeting and planning process. However, some counties such as county D did not fully adopt the process, especially in sub-county hospitals.

#### Frontline workers were involved in the planning process during the development of the Annual Work Plan (AWP) but they were not involved in developing the PBB

In all four counties, the county health management team (CHMT) made all the decisions regarding budgets. The sub-county health management teams and the facility in-charges were left out of the budgeting process.

> *“It is a problem because we are the key makers concerning budget. We must start from down going up. But often, I don’t know if what I budgeted for was captured on the major budget. Is there a match or a mismatch on it, I can’t tell”? Facility Manager County B*

Failure to involve the frontline workers was felt to affect health system efficiency in various ways. First, most facilities had never seen the county budget and had no idea what was being implemented in the county. This limited accountability at the facility and county level.

> *“they’ll procure drugs and non-pharmaceuticals worth say 90,000,000 for the whole county. But you can’t know exactly how much was spent on your facility. You can estimate based on the drawing rights but you can’t be 100% sure that this is the money that was spent on my facility*.*” Facility Health Manager, County A*

Second, it was felt that because facilities were unclear about what was being implemented, they were unable to effectively evaluate their achievements against budgets.

> *“We can evaluate revenue from NHIF that we collect and spend at source and put in place measures to improve performance. But it’s hard to evaluate what the county probably spends on us” Facility Health Manager, County A*

Third, counties ended up making decisions that were not consistent with the needs of the health facilities. Blanket decisions wasted resources. For example, in county A, the department was not able to access materials for the orthopedic department because it was not something that was commonly used in the county, yet for their facility, this was an important need. The facility had an orthopedic surgeon whose skills would be wasted if these materials were not availed.

> *“They need to involve us, otherwise they waste resources. They should ask us, what do you need? What is not important? what will improve your performance? Other than just pushing, because yes, you’ve bought me drugs worth 2 million drugs, but what if my catchment population does not match the drugs that you bought me?” Facility Health Manager, County C*

Overall, both public and facility involvement was said to improve transparency. Improved transparency enabled efficiency by enhancing accountability for results.

> *“To me, I think facility and public involvement is the best way of practice for accountability and transparency, that’s the best way to become accountable. Whatever we are doing, everybody needs to be at par, they should know what is happening within the system*.*” Facility Manager County B*

### Priority setting and resource allocation criteria

The priority setting process was not evidence-based. Participants noted that priorities for the recurrent budget were decided based on historical expenditure and lobbying, while priorities for development budget were set based on demands from political leaders which had a bias towards visible infrastructure expenditure that enhanced positive citizen perception about their performance as politicians. For example, it was reported that politicians prioritized the construction of new health facilities without considering the population demand for health services. As a result, the CDOH had developed more health facilities than were needed to meet the population demand for facility-based health services and stretched the other limited resources such as human resource for health.

> *“They [MCAs] say they want health centers. Right now, I’ve got about 30 health facilities which are complete. But, I’m not able to equip or staff. Every MCA wants to tell the electorate I have built this one, but why have a facility that is not functional? I hope as we mature in devolution, they’ll understand that facilities should be constructed, in accordance with the population and the county’s ability to staff and to equip them” County Health Manager County B*

Second, the need to maximize revenue meant that the income generating departments were given priority over other departments. For example, in county A, hospitals had access to only one source of fund - reimbursement for maternal deliveries dubbed “Linda mama”. As this was their only source of funds, they prioritized the allocation of budgets to the maternity department to raise more revenue. In some cases, this happened at the expense of other health system needs and goals.

> *“For Linda mama funds, priorities mainly goes towards the maternal care, their priorities are considered first. If they want a baby and mother package, we must budget for that. Basically, they have more power to decide what they want to do within the department. The other departments only benefit if they support maternity”* Facility Health Manager County A

### Budget approval process

#### Budgets were reprioritized during approval

As the budget went through the county treasury, the county executive, and the assembly, some health priorities were changed without the knowledge of the county department of health. Respondents noted that the changes further misaligned the budget from the plans. While the county executive made changes to the budget, these changes were not reflected in the developed plans. Second, in county C, the county assembly reduced the resources available to the CDOH in the budget approval process further exacerbating the resource challenges.

> *“The budget has been changing without us knowing. Whatever goes to the assembly is not what we get back. For long, we thought it was the assembly making the changes. But we discussed with the assembly last year and they told us the changes come from the executive” County Health Manager County B*
>
> *“…And the budget goes to the county treasury then they, I don’t know what they do if we budget, they reduce our budget and even the one, which is reduced is nowhere to be seen” County Health Manager County C*

## Discussion

This study examined how the budget formulation process affects health system efficiency at the county level in Kenya. We report several weaknesses across all the 6 aspects of the budget formulation process that have potential implications for county health system efficiency. First, we find that in two counties, budget ceilings were determined using historical allocations. In addition, ceilings were either late or not forthcoming, insufficient, and not cascaded to lower-level priority healthcare facilities. These findings resonate with findings from Ghana and Democratic Republic of Congo (DRC) where ceilings were not indicative^19,20^ and Thailand’s civil servant scheme where ceilings were allocated historically ^21^. The historical allocation of budgets undermines health system efficiency by ignoring the evolution of health sector priorities. When ceilings are not provided or provided late, it renders the budgeting process moot since budgets are not aligned with the reality of resource availability. This disempowers health sector units (county department of health, health facilities) from effectively contributing to the budgeting process with the implication that budgets will not be aligned with actual health sector needs, thus compromising the optimal use of resources. When ceilings are insufficient, they compromise efficiency by constraining health system investments and hence health system input mix with negative implications for health outcomes. The failure to cascade budget ceilings to peripheral healthcare facilities had the same effect: disempowering these budgeting units from contributing to the budgeting process. Given that peripheral healthcare facilities predominantly provide primary healthcare (PHC), their disempowerment in the budgeting process undermines PHC delivery, against a backdrop where PHC has shown to be cost-effective and hence efficiency enhancing ^22^.

Second, on budget structure, we found that while on paper counties are supposed to use programme-based budgets, in practice, the budgets are still line-item based. The use of line-item budgets led to budget rigidities which limited the capacity of counties to respond to emergent healthcare needs. Further, we found that budgets were not aligned to plans. This echoes previous findings in Kenya where there was institutionalized misalignment of the planning and budgeting processes at the national level ^23^. Similar findings have also been reported in Lesotho where budgets and plans were misaligned as they happened under different structures ^24^. The misalignment between budget and plans meant that budgets did not adequately represent health sector priorities, which could compromise allocative efficiency.

Third, the limited involvement of front-line service providers and the community in the development of county health sector budgets and plans led to the misalignment of budgets with population health priorities and limited budget accountability, with implications for both technical and allocative efficiency. This limited involvement of frontline providers and the community has also been documented in previous findings in Kenya where decentralization resulted in limited hospital autonomy ^8^.

Fourth, we found that the persistence of off-budget donor funding compromised health sector planning because CDoHS were unaware of the total available resources in the health sector. This in turn led to duplication of efforts which compromised the technical efficiency of county governments. This finding is like findings in Sierra Leone where there were multiple funding sources with multiple bank accounts limiting transparency ^16^. We also found that health sector funding at the county level was fragmented. This is similar to China where there was fragmented financing of the health system by the government at the national and sub national level ^25 4^. Fragmented funding limited effective planning and budgeting by limiting pooling of resources and shifting decision making over resources to entities outside the health sector.

Fifth, we found that the budget formulation processes were dominated by informal priority setting criteria such as lobbying. This compromised allocative efficiency of health systems by compromising the optimal allocation of heath sector resources ^10,21,26^.

Lastly, we found that CDoH autonomy was usurped during the budget approval process where the county treasury and county assembly often revised the budgets without reference to the county department of health. The reprioritization of budget by the county assembly at the approval stage without reference to the county department of health and its stakeholder holders disempowered health sector stakeholders. This had the potential of misaligning final budgets with health sector priorities and hence compromising both technical and allocative efficiency ^4^.

While most budget formulation challenges such as use of line budgets cut across both efficient and inefficient counties, some aspects were different. County A, one of the best performing counties reported more participation of frontline health workers in the budgeting process and better relationships with off budget donors. County B, another well performing county had the highest allocation to the health sector, and moderate involvement of the CDOH in planning and budgeting. County C one of the poor performing counties had significant challenges across all the areas of the budget process beginning with complete failure to disclose ceilings. County D, another poor performing also had poor involvement of the CDOH. The findings on PFM practices were therefore mixed rather than systematically different between the counties that were ranked as efficient and the ones that were ranked as inefficient by the quantitative efficiency analysis. This would be because the nature of PFM practices documented are perverse in Kenyan counties, with differences in degrees across countries that are difficult to tease out using a qualitative approach. It could also be because the counties that were ranked as efficient by the quantitative analysis by being on the efficiency frontier are inefficient in absolute terms, even though they are relatively more efficient than the counties that are at a distance from the efficiency frontier.

A limitation of this study is the inclusion of only four out of the 47 counties in Kenya. This is important because budget formulation practices vary across counties. However, as is characteristic of qualitative case studies, the intention was not to achieve statistical generalization but rather analytical generalization ^27^. A second limitation is the different financial data reported by the government agencies dealing with the county finance. Reports from the county differ from those by the controller of budget which also differ from those reported by the national treasury. We used the most complete data available.

Despite the limitations, the meta issues identified within this study are analytically generalizable and we can draw various recommendations from the findings. First, the county treasury should use the MTEF reports from the county departments as a guide to develop final budget ceilings. MTEF reports outline the requests of the departments and the public for the health system over a 3year period. Second, the county finance departments should provide timely ceilings to allow sufficient time for PBB development. The budget process guidelines state that the CFSP containing the budget ceilings should be approved and disseminated by 14^th^ March of the planning year. Third, counties should allocate enough resources to the health sector. The intergovernmental collaboration agreement recommends a minimum of 30% of the total county budget allocation to the CDOH. Fourth, the CDOHs should fully cascade the budget ceilings to the different planning units and allow each unit to make its own budget as guided by the county financing guidelines. Fifth, while the counties develop many budget formats, they should use the programme-based budget as the functional budget as required by the PFMA. Sixth, the CDOHS should develop the PBB to completion, and ensure that the plans and budgets align. Seventh, health sector stakeholders should pool all health sector funds from different sources to enhance efficiency. This can be achieved through empowering the forums created to strengthen joint planning and budgeting. These include the County Health Stakeholders Forums and the Health Sector Working Groups. Eighth, public participation should be better structured and guided to ensure that the public makes informed decisions. Ninth, health workers who are most aware of health system needs should be at the center of the decision-making process in the health budgeting and planning process. This can be achieved through strengthening the health facility boards and committees. Tenth, priorities in the planning and budgeting decisions should be evidence informed. This will ensure resources are allocated to high impact interventions. Finally, county assembly involvement in the budget process should be limited to oversight as is required by the legal framework.

## Conclusion

This study examined the relationship between budget formulation and efficiency of county health systems in Kenya. A key finding is that the budget formulation process influences both technical and allocative efficiency directly or by influencing the implementation and/or evaluation of the budget. A well formulated budget is therefore important in ensuring a well implemented and evaluated budget which will in turn reduce inefficiencies. By enhancing the budget formulation process, the health system will get more health from the available resources. This study has highlighted six aspects of the budget formulation process that ought to be strengthened to enhance efficiency; budget ceilings, budget structure, participatory budget formulation, pooling of health funds, priority setting processes and the budget approval process.

## Data Availability

All data produced in the present study are available upon reasonable request to the authors

## Acknowledgements

This work was funded by a MRC/DFID/Wellcome Trust Health Systems Research Initiative (HSRI) grant no MR/R01373X/1. This work is published with the permission of the Director of KEMRI.

